# Associations of Water, Sanitation, and Hygiene with Typhoid Fever in Case-Control Studies: A Systematic Review and Meta-Analysis

**DOI:** 10.1101/2022.07.22.22277917

**Authors:** Chaelin Kim, Gerard R Goucher, Birkneh Tilahun Tadesse, Woojoo Lee, Kaja Abbas, Jong-Hoon Kim

**Affiliations:** International Vaccine Institute, Seoul, South Korea; Seoul National University, Seoul, South Korea; London School of Hygiene & Tropical Medicine, London, United Kingdom

## Abstract

**Background:** Typhoid fever is a major public health issue in low- and middle-income countries. It is transmitted through fecally contaminated food or water and thus improving water, sanitation, and hygiene (WASH) is considered key to its control. We sought to quantify the association between WASH and typhoid fever.

**Methods:** We updated a previous review by including new findings from the literature indexed in Web of Science, Embase, and PubMed. We kept the search terms, typhoid and case-control, consistent with the previous review. We assessed the risk of bias using the Risk of Bias in Non-Randomized Studies of Interventions (ROBINS-I). We categorized WASH exposures according to the WHO/UNICEF Joint Monitoring Programme for Water Supply, Sanitation and Hygiene (JMP) classification. We conducted a Bayesian random-effects meta-analysis of odds ratios (ORs) extracted from the studies without a critical risk of bias.

**Findings:** We identified 25 eligible articles including 19 articles from the previous review. Pooled ORs indicated limited hygiene (OR = 2.26, 95% CrI: 1.38 to 3.64), untreated water (OR = 2.21, 95% CrI: 1.53 to 3.48) and using surface water (OR = 2.16, 95% CrI: 1.24 to 3.60) increased odds of culture-confirmed typhoid fever. On the other hand, basic hygiene (OR = 0.6, 95% CrI: 0.38 to 0.97) and treated water (OR = 0.62, 95% CrI: 0.41 to 0.89) reduced odds of culture-confirmed typhoid fever.

**Conclusion:** Our analyses updated quantitative evidence of association between WASH and typhoid fever. Our study findings will be useful to infer actionable insights on the most effective ways to control typhoid fever in low- and middle-income countries. Our analyses also offer a possibility to leverage JMP WASH data to explore potential burden of typhoid fever.

**Systematic review registration:** PROSPERO 2021 CRD42021271881

**Author Summary:** Typhoid fever is a major public health issue in the low- and middle-income countries. It is transmitted through fecally contaminated food or water and thus improving water, sanitation, and hygiene (WASH) is considered key to its control. We quantified the association between WASH and typhoid fever through a systematic review of the case-control studies and meta-analyses of extracted odds ratios (ORs). We categorized WASH exposures according to the WHO/UNICEF Joint Monitoring Programme for Water Supply, Sanitation and Hygiene (JMP). We used a Bayesian random-effects model to account for the heterogeneity of studies that were conducted at different times and places, and adjusted potential bias differently. Pooled ORs indicated that limited hygiene (OR = 2.26, 95% CrI: 1.38 to 3.64), untreated water (OR = 2.21, 95% CrI: 1.53 to 3.48) and using surface water (OR = 2.16, 95% CrI: 1.24 to 3.60) increased odds of typhoid fever. On the other hand, basic hygiene (OR = 0.6, 95% CrI: 0.38 to 0.97) and treated water (OR = 0.62, 95% CrI: 0.41 to 0.89) reduced odds of culture-confirmed typhoid fever. Our analyses updated evidence for the association between WASH and typhoid fever. The updated evidence strongly supports that improved WASH such as improved water source, water treatment, and basic hygiene will help reduce typhoid fever in low- and middle-income countries. By linking WASH exposures to new JMP WASH categories our analyses also offer a possibility to leverage JMP WASH data sets to explore potential burden of typhoid fever.

## Introduction

Typhoid fever, an infection caused by *Salmonella enterica* serovar Typhi (*S*. Typhi), is a global public health problem. An estimated 11 to 20 million typhoid fever cases including 128,000 to 161,000 deaths occur each year (1–4) with the majority in low- and middle-income countries (LMICs) (5,6). Although several effective treatment and prevention strategies are available (7), improving water, sanitation, and hygiene (WASH) is considered key to preventing typhoid fever considering that *S*. Typhi is transmitted via fecally contaminated water or food (8).

Understanding the relative strengths of the association between different components of WASH and typhoid fever may lead to more cost-effective strategies for implementing various WASH components that can provide the strongest protection against typhoid fever. Designing such a strategy requires a detailed understanding of the strength of the association between different components of WASH and typhoid fever.

Population levels of access to improved WASH are monitored by the WHO/UNICEF Joint Monitoring Programme for Water Supply, Sanitation and Hygiene (JMP) in over 190 countries since 1990 (9). The JMP WASH classification has three categories–drinking water, sanitation, and hygiene–and each category has service ladders indicating different levels of improvement. For instance, the drinking water category has five service ladders: Safely managed, Basic, Limited, Improved, Unimproved, and Surface water. JMP maintains estimates on each of the different categories that can be compared across each of the 190 countries that cover almost all of the LMICs.

Understanding the strength of the association between the levels of WASH and typhoid fever risk can create an opportunity to leverage the efforts of the JMP to better understand the risk of typhoid fever within and across countries. While there appears to be a clear association between typhoid fever and the levels of WASH practices, the degree of association varies from study to study. The systematic review and meta-analysis by Mogasale et al. (10) summarized the findings from case-control studies on the association between the levels of WASH and typhoid fever. This study focused only on the drinking water source and exposure categories of the included studies were not classified according to the JMP WASH categories. The systematic review and meta-analysis by Brockett et al. (11) included all three categories of WASH and categorized WASH exposures from case-control studies according to JMP WASH classification, but was applied in a broader level without using specific service ladders. Both studies included findings based on Widal-confirmed typhoid fever cases in addition to cases confirmed through blood culture, which may introduce bias because of the low specificity of the Widal test (12).

In this study, we aim to improve the estimates for the association between WASH exposures and typhoid fever by including new findings published since the previous review done by Brockett et al. (11), applying a more rigorous risk of bias assessment, and clarifying the association between the JMP WASH categories and WASH exposures measured in case-control studies. Our study findings will be useful to infer actionable insights on the most effective ways to prevent the spread of typhoid fever and the ways to leverage the WHO/UNICEF JMP WASH data to explore the potential burden of typhoid fever.

## Methods

### Search strategy

We searched three databases – Web of Science, Embase, and PubMed – to find peer-reviewed articles in English. In each database, the title and abstract of the paper were searched using the following search terms: (“case control” OR “case-control”) AND “typhoid”. The search terms were consistent with the previous review except that we did not include “retrospective” to restrict our search to case-control studies. We restricted our search to articles published during June 2018 - July 2021 to identify articles that were published after the publication of Brockett *et al*. study (11), which included articles published between January 1990 and June 2018.

### Inclusion and exclusion criteria

Inclusion and exclusion criteria were developed based on the population, intervention, comparison, outcomes, and study design (PICOS) framework (13). These predefined criteria were included in the protocol published in PROSPERO (14). Eligible study populations encompass populations of all ages, genders, and socioeconomic statuses living in low- and middle-income countries. Studies would be eligible for inclusion if they considered one of five WASH exposure categories, specifically: water source, water management, water treatment, sanitation, and hygiene. Preventive exposures mitigate typhoid fever transmission, and risk exposures increase typhoid fever transmission. We excluded studies that were meant to evaluate vaccine efficacy in which the nature of interactions between WASH exposures and vaccination is not clear. Studies were considered eligible if they investigated association between typhoid fever and at least one WASH exposure using an odds ratio (OR).

### WASH exposure categories

Studies vary in their WASH exposures, and we tried to systematically map the WASH exposures from included studies to the JMP WASH categories and service ladders (**Table 1**). The JMP provides service ladders for each of the three WASH categories: drinking water, sanitation, and hygiene. In addition to these three categories, we used two additional categories of water treatment and water management to delve into other important characteristics of water interventions. These two categories were also used in the previous review by Brockett *et al*. Food hygiene was not covered in our study as we used the JMP service ladder for hygiene category which focuses on handwashing.

**Table 1.** WASH-related exposures from included studies and corresponding WASH service ladders. The following table includes the WASH category, service ladders and examples of WASH exposures. This categorization was used to classify the extracted data for the meta-analysis.

| WASH category | Service ladders | Example WASH exposures from included studies |
| --- | --- | --- |
| Water source<sup>††</sup> | Improved (Safely managed, Basic, Limited)* | Drinking piped water only, tube well water, etc. |
|  | Unimproved | NA |
|  | Surface water | Drinking river water, surface water, stream water, unboiled surface water, etc. |
| Sanitation | Safely managed | NA |
|  | Basic | NA |
|  | Limited | NA |
|  | Unimproved | Unimproved pit latrine |
|  | Open defecation | Places used to defecate (field, pond, river, canal, nearby stream), sewage disposal directly to the environment etc. |
| Hygiene | Basic | Use of soap for handwashing, soap hand wash before food/after defecation/after urination, soap available to wash hands, soap observed in home. |
|  | Limited | Soap not available near toilet, no use of soap for handwashing. |
|  | No facility | NA |
| Water treatment<sup>†</sup> | Treated water | Drinking purified water, boiled water in home, treated water in home, disinfected water at home using boiling or filtration. |
|  | Untreated water | Drinking untreated water, drinking unboiled water, grossly contaminated home water. |
| Water management<sup>†</sup> | Safe water storage | Storage of water in covered container, narrow-mouth container, metal covering, wide mouthed container with lid, a narrow mouthed container (with lid). |
|  | Unsafe water storage | Dirty container for storing drinking water. |

\* While JMP WASH classification provides five service ladders, WASH exposures from individual studies do not provide sufficient descriptions to match any of the three service ladders. We grouped the three ladders into a single category called improved water source. The definition of “improved” is also defined by the JMP (9).
<sup>†</sup> These categories are not part of the JMP WASH classification.
<sup>††</sup> The original category name is “drinking water” under the JMP classification.
NA - not available

Specific WASH exposures from included studies were first checked to see if they matched the JMP ladder definitions. If they matched one of these definitions, the exposure would be placed into the corresponding JMP ladder. For instance, basic in the JMP hygiene ladder is defined as “availability of a handwashing facility with soap and water at home”. Relevant exposures such as the use of soap for handwashing or soap available to wash hands are classified under the basic hygiene category. The five WASH categories with 15 subcategories were used in our study to synthesize the findings on the association between the WASH characteristics and typhoid fever.

### Data extraction

Two independent reviewers (GG, JHK) determined the eligibility of articles in two separate phases. Any disagreements were resolved by a third reviewer (CK). Initially, titles and abstracts were screened to ensure that the studies used the case-control methodology, that the outcomes are typhoid cases, and that the context was in LMIC. Then, full manuscripts were read to ensure that articles met all of our PICOS criteria. Two reviewers (GG, CK) extracted data from the included studies, including author information, publication year, case/control definitions, WASH exposures, diagnostic methods, country, and effect size (odds ratio) for individual exposures. Google Sheets was used to manage the data.

### Risk of bias assessment

We assessed the risk of bias of the included studies using the Cochrane Risk of Bias in Non-Randomized Studies of Interventions (ROBINS-I) tool (15) in seven domains: 1) confounding, 2) selection, 3) intervention classification, 4) intervention deviation, 5) missing data, 6) outcome measurement, and 7) selective reporting. Based on the assessment results in each domain, the studies were labeled as having a low, moderate, serious, or critical risk of bias. Two authors (CK, JHK) examined the risk of bias independently, and any discrepancies were resolved by discussion.

### Statistical analysis

Data from studies that were expected not to have the critical risk of bias were used to generate the pooled estimates. Studies that did not use culture-confirmed cases were not included in any data synthesis. The analyses were performed using the R statistical software (version 4.1.3). We developed a series of Bayesian random effects models using the *brms* package (16) to estimate the pooled ORs with 95% credible intervals (CrIs) for each exposure category with more than two studies. Random effects models were utilized as we assume that true effects may vary for each study depending on the contexts. Bayesian meta-analyses are particularly useful when the number of studies is small and enable us to use prior knowledge (17). We assessed the possibility of publication bias through visual inspection of the funnel plots (**Appendix B**). The repository for the data and software code of this study are publicly accessible at the GitHub repository (18).

## Results

### Characteristics of included studies

The PRISMA flow diagram in **Figure 1** depicts the different phases of a systematic review. The search yielded 14, 26, and 38 articles from Web of Science, PubMed, and Embase, respectively. We obtained 51 unique articles after removing the duplicates. After reviewing the title and abstract, we excluded 43 non-eligible articles and reviewed the full-text copies of 8 studies. Following the full-text review, 5 new studies were included in our review in addition to the 19 studies included in the previous review conducted by Brockett et al. (11). Thereby, 24 studies were included in our review. The newly identified studies are from the Democratic Republic of Congo, Fiji, Malawi, Pakistan and Uganda (19–23). Among the 24 included studies, blood culture was used to fully define cases in 10 studies (42%) and define the subset of cases in 5 studies (21%). **Table 2** contains information on the characteristics of the included studies. After removing the studies with critical risk of bias, the 15 studies were included in the meta-analysis (**Figure 1**). We performed a meta-analysis for the 7 categories for which there were more than two studies. Summaries of the meta-analysis results are shown in **Table 3**.

**Table 2.** Characteristics of studies. Characteristics of included studies in the systematic review, and information on the JMP WASH category, study, country, exposures, effect size (odds ratio or adjusted odds ratio), and diagnostic methods.

| JMP WASH Category | Study | Country | Exposures | Effect Size (Odds Ratio) | Diagnostic Methods |
| --- | --- | --- | --- | --- | --- |
| Water Source (Improved) | Bhuniah et al. 2009 (8) | India | Drinking water (Piped water only) | 7.3 (2.5-21) | Widal test |
|  | Bhuniah et al. 2009 (8) | India | Drinking water (Tube well water) | 0.25 (0.08-0.75) | Widal test |
|  | Ram et al. 2007 (24) | Bangladesh | Pipe to municipal supply | 0.5 (0.2-1.1)<sup>†</sup> | Blood culture |
|  | Sharma et al. 2009 (25) | India | Drinking water (Piped water supply at home) | 0.4 (0.2-0.9) | Widal test |
|  | Srikantiah et al. 2007 (26) | Uzbekistan | Drinking water habits outside home (Piped water) | 0.6 (0.3-1.6)<sup>†</sup> | Blood culture |
|  | Vollaard et al. 2004 (27) | Indonesia | Drinking water (Piped water) | 0.44 (0.19-1.01)<sup>†</sup> | Blood culture |
| Water Source (Surface water) | Aye et al. 2004 (28) | Myanmar | Drinking untreated river water | 12.5 (2.8-75.3)* | Diazo urine test or positive Widal test |
|  | Luxemburger et al. 2001 (29) | Vietnam | Drinking river water | 1.8 (0.8-5.6)<sup>†</sup> | Blood culture |
|  | Prasad et al. 2018 (21) | Fiji | Drinking from an alternative water source (surface water source) | 3.61 (1.44-9.06)*<sup>†</sup> | Blood culture |
|  | Prasad et al. 2018 (21) | Fiji | Main household water source (Surface water source) | 1.28 (0.35-4.70)<sup>†</sup> | Blood culture |
|  | Sharma et al. 2009 (25) | India | Drinking water (Stream water at home) | 1.6 (0.9-2.6) | Widal test |
|  | Srikantiah et al. 2007 (26) | Uzbekistan | Drinking unboiled surface water outside home | 3 (1.1-8.2)*<sup>†</sup> | Blood culture |
|  | Srikantiah et al. 2007 (26) | Uzbekistan | Surface water at home | 1.9 (0.7-4.9)<sup>†</sup> | Blood culture |
| Water Treatment (Treated water) | Bhuniah et al. 2009 (8) | India | Drinking water (Purification) | 0.44 (0.19-1.0) | Widal test |
|  | Luby et al. 1998 (30) | Pakistan | Home water (Clean home drinking water) | 0.9 (0.4-1.9)<sup>†</sup> | Blood culture |
|  | Mermin et al. 1999 (31) | Tajikistan | Boiled water in home | 0.2 (0.05-0.6)*<sup>†</sup> | Blood and stool culture |
|  | Mirembe et al. 2019 (23) | Uganda | Do you treat your drinking water? | 9.23 (4.84-17.61)* | Diagnosis of clinical signs |
|  | Muti et al. 2014 (32) | Zimbabwe | Boil drinking water | 0.24 (0.07-0.90)* | Diagnosis of clinical signs |
|  | Prasad et al. 2018 (21) | Fiji | Treated water in house | 0.89 (0.57-1.39)<sup>†</sup> | Blood culture |
|  | Ram et al. 2007 (24) | Bangladesh | Disinfected water at home using boiling or filtration | 0.7 (0.4-1.6)<sup>†</sup> | Blood culture |
|  | Sharma et al. 2009 (25) | India | Drinking water (Drinking boiled water) | 1.3 (0.6-2.6) | Widal test |
|  | Srikantiah et al. 2007 (26) | Uzbekistan | Drinking water habits outside home (Boiled water) | 0.5 (0.3-0.9)<sup>†</sup> | Blood culture |
|  | Srikantiah et al. 2007 (26) | Uzbekistan | Drinking water habits at home (Boiled water) | 0.4 (0.2-0.8)<sup>†</sup> | Blood culture |
| Water Treatment (Untreated water) | Alba et al. 2016 (33) | Indonesia | Water treatment before drinking (Never) | 1.68 (0.99-2.82)<sup>†</sup> | Serology and culture |
|  | Luby et al. 1998 (30) | Pakistan | Home water (Grossly contaminated) | 1.1 (0.6-2.1)<sup>†</sup> | Blood culture |
|  | Luxemburger et al. 2001 (29) | Vietnam | Drinking unboiled water | 1.5 (0.8-3.3)<sup>†</sup> | Blood culture |
|  | Mermin et al. 1999 (31) | Tajikistan | Drinking unboiled water | 9.6 (3.0-34.0)*<sup>†</sup> | Blood and stool culture |
|  | Prasad et al. 2018 (21) | Fiji | Drinking untreated water | 1.8 (1.07-3.03)<sup>†</sup> | Blood culture |
|  | Ram et al. 2007 (24) | Bangladesh | Drinking unboiled water at home | 12.1 (2.2–65.6)*<sup>†</sup> | Blood culture |
|  | Srikantiah et al. 2007 (26) | Uzbekistan | Drinking water habits outside home (Unboiled water) | 3 (1.5-6)<sup>†</sup> | Blood culture |
|  | Srikantiah et al. 2007 (26) | Uzbekistan | Drinking water habits at home (Unboiled water) | 2.1 (1-4.4)<sup>†</sup> | Blood culture |
|  | Tran et al. 2005 (34) | Vietnam | Drinking untreated water | 3.9 (2.0-7.5)*<sup>†</sup> | Blood or stool culture |
| Water Management (Safe water storage) | Bhunias et al. 2009 (8) | India | Drinking water (Covered container) | 0.25 (0.08-0.75) | Widal test |
|  | Bhunias et al. 2009 (8) | India | Drinking water (Narrow mouth container) | 0.35 (0.15-0.76) | Widal test |
|  | Karkey et al. 2013 (35) | Nepal | Metal covering of water storage | 0.22 (0.1-0.6) | Blood culture |
|  | Muti et al. 2014 (32) | Zimbabwe | Store water in wide mouthed container with lid | 3.68 (1.62-8.35)* | Diagnosis of clinical signs |
|  | Muti et al. 2014 (32) | Zimbabwe | Storage of water in a narrow mouthed container with lid | 0.43 (0.22-0.8)* | Diagnosis of clinical signs |
|  | Sharma et al. 2009 (25) | India | Storage of water (Narrow-mouthed container) | 0.4 (0.2-0.7) | Widal test |
|  | Srikantiah et al. 2007 (26) | Uzbekistan | Storing water (Keep water container covered) | 0.2 (0.04-1.1) | Blood culture |
| Water Management<br>(Unsafe water storage) | Bhan et al. 2002 (36) | India | Dirty container for storing drinking water | 1.99 (0.6-6.65)* | Blood culture |
| Sanitation<br>(Unimproved) | Prasad et al. 2018 (21) | Fiji | Unimproved pit latrine | 49.47 (9.42–259.92)* | Blood culture |
| Sanitation<br>(Open defecation) | Alba et al. 2016 (33) | Indonesia | Places used to defecate (Field) | 0.98 (0.72-1.34)<sup>†</sup> | Serology and culture |
|  | Alba et al. 2016 (33) | Indonesia | Places used to defecate (Pond/river/canal) | 0.77 (0.56-1.06)<sup>†</sup> | Serology and culture |
|  | Luxemburger et al. 2001 (29) | Vietnam | Defecation in a fish pond or river | 1.1 (0.5-3.2)<sup>†</sup> | Blood culture |
|  | Prasad et al. 2018 (21) | Fiji | No toilet (Open defecation) | 9.87 (0.85–114.35)*<sup>†</sup> | Blood culture |
|  | Sharma et al. 2009 (25) | India | Toilet facilities (Nearby stream) | 1.5 (0.9-2.7) | Widal test |
|  | Tran et al. 2005 (34) | Vietnam | Sewage disposal directly to the environment | 2.4 (1-5.8)<sup>†</sup> | Blood or stool culture |
| Hygiene (Basic) | Aye et al. 2004 (28) | Myanmar | Hand washing with soap | 0.15 (0.03-0.81)* | Diazo urine test or positive Widal test |
|  | Bhunja et al. 2009 (8) | India | Soap hand wash before food | 1.9 (0.8-4.4) | Widal test |
|  | Bhunja et al. 2009 (8) | India | Soap hand wash after defecation | 0.3 (0.12-0.75) | Widal test |
|  | Bhunja et al. 2009 (8) | India | Soap hand wash after urination | 0.08 (0.03-0.26) | Widal test |
|  | Gauld et al. 2019 (20) | Malawi | Soap available to wash hands after toilet use in the previous 3 weeks | 0.6 (0.4-.98)*<sup>†</sup> | Blood culture |
|  | Mirembe et al. 2019 (23) | Uganda | Do you wash hands with soap? | 1.81 (0.95–3.45)* | Diagnosis of clinical signs |
|  | Prasad et al. 2018 (21) | Fiji | Use soap for handwashing | 0.61 (0.37–0.95)*<sup>†</sup> | Blood culture |
|  | Ram et al. 2007 (24) | Bangladesh | Soap observed in home | 0.5 (0.2-1.3)<sup>†</sup> | Blood culture |
| Hygiene (Limited) | Alba et al. 2016 (33) | Indonesia | Soap near toilet (Never) | 4.4 (2.0-9.65)<sup>†</sup> | Serology and culture |
|  | Bhan et al. 2002 (36) | India | Nonuse of soap for washing hands | 1.82 (1.04-3.21)*<sup>†</sup> | Blood culture |
|  | Siddiqui et al. 2008 (37) | Pakistan | Soap available near hand washing facility (No) | 2.6 (1.1-6.3)*<sup>†</sup> | Blood culture or serology test |
|  | Velema et al. 1997 (38) | Indonesia | Does not use soap when washing hands | 29.8 (2.19-407.0)* | Diagnosis of clinical signs and Widal test |
|  | Vollaard et al. 2004 (27) | Indonesia | No use of soap for handwashing | 1.91 (1.06-3.46)*<sup>†</sup> | Blood culture |

\* adjusted odds ratio from multivariate analysis
<sup>†</sup> these values were included in the meta-analysis

**Table 3.** Meta-analyses of WASH exposures and typhoid fever. The results of the meta-analyses of WASH exposures and typhoid fever are shown with pooled odds ratio and 95% credible intervals. Standard deviation (SD) of true effects represents τ value. The results were also compared with the previous studies of Brockett et al (11) and Mogasale et al (10).

| WASH indicators | *N* * | Pooled OR<br>[95% CrI] | SD of true<br>effects | Brockett<br>(2020) | Mogasale<br>(2018) |
| --- | --- | --- | --- | --- | --- |
| Improved water source | 4 | 0.54 [0.31, 1.08] | 0.29 | 0.73 [0.56, 0.95] | 0.70 [0.46, 1.05] |
| Treated Water | 6 | 0.62 [0.41, 0.89] | 0.29 | 0.59 [0.45, 0.75] | NA |
| Basic hygiene | 3 | 0.60 [0.38, 0.97] | 0.24 | 0.52 [0.40, 0.67] | NA |
| Surface water | 5 | 2.16 [1.24, 3.60] | 0.27 | 1.85 [1.37, 2.49] | NA |
| Untreated water | 9 | 2.21 [1.53, 3.48] | 0.42 | 2.39 [1.95, 2.93] | NA |
| Open defecation | 5 | 1.06 [0.71, 2.10] | 0.39 | 0.99 [0.84, 1.18] | NA |
| Limited hygiene | 4 | 2.26 [1.38, 3.64] | 0.29 | 2.20 [1.86, 2.60] | NA |

\*Number of exposures
NA - not available

**Figure 1.**
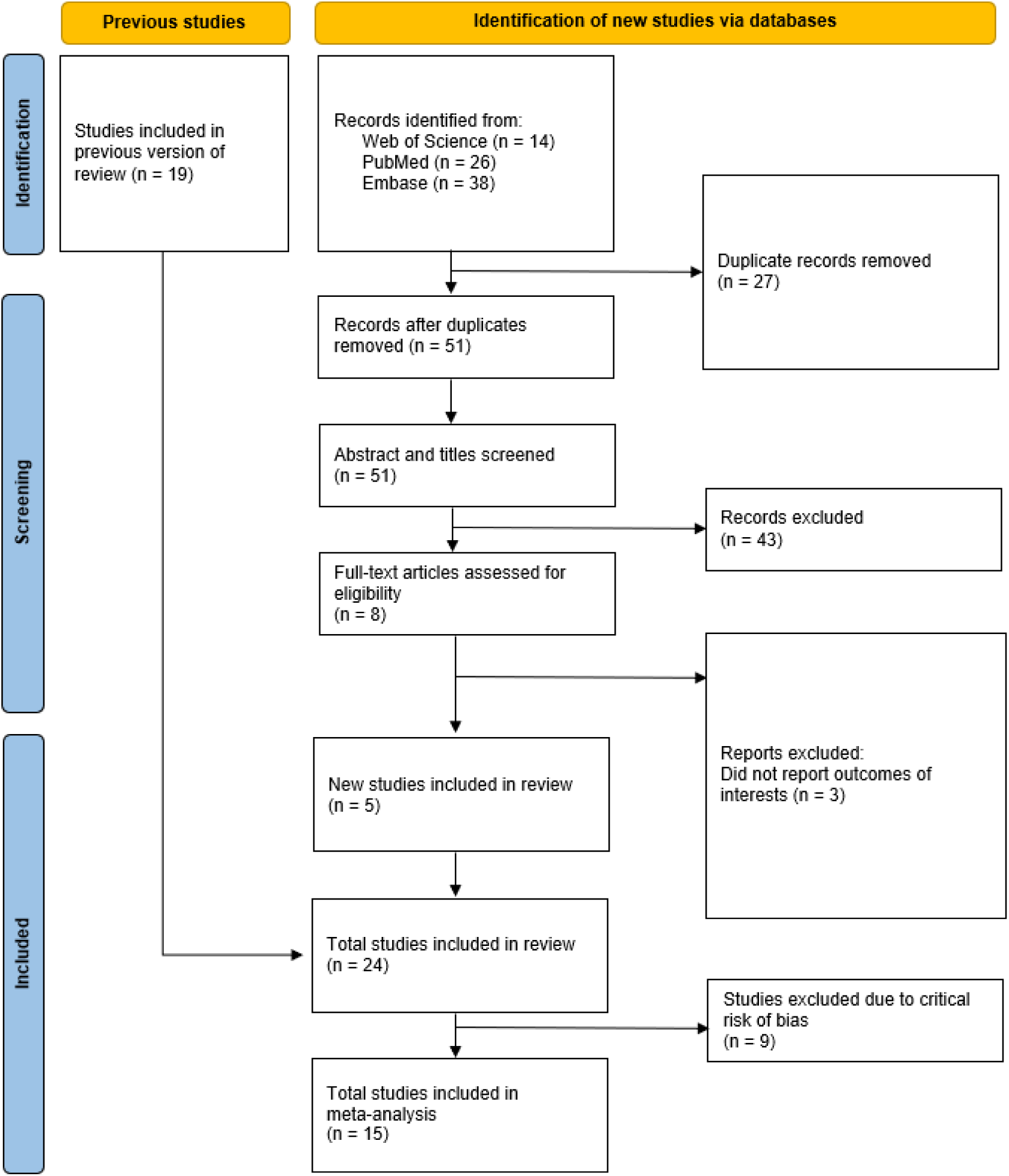
PRISMA flow diagram. The PRISMA (Preferred Reporting Items for Systematic Reviews and Meta-Analyses) flow diagram shows the number of articles at the different phases of identification, screening, and inclusion in the systematic review and meta-analysis.

### Risk of bias assessment

Except for five studies that were categorized as having an overall moderate risk of bias, all other studies were classified as having the overall serious or critical risk of bias since at least one domain was assessed as having the serious risk of bias (**Figure 2**). The majority of included studies were graded as having a moderate risk of bias in the domain of confounding as the known important confounding domains (age, sex, and socioeconomic characteristics) were adequately measured and controlled even though some level of confounding is still expected due to the nature of the case-control study. In the domains of intervention classification, deviations from intended interventions, and the selection of the reported result, the majority of included studies were also classified as having a moderate risk of bias. In addition, ten studies were labeled as having a low risk of bias as they utilized a culture-confirmed typhoid fever diagnosis. However, most studies were rated as having a serious risk of bias as the case-control research design is prone to selection bias. Lastly, ten studies did not provide adequate information to assess bias due to missing data.

**Figure 2.**
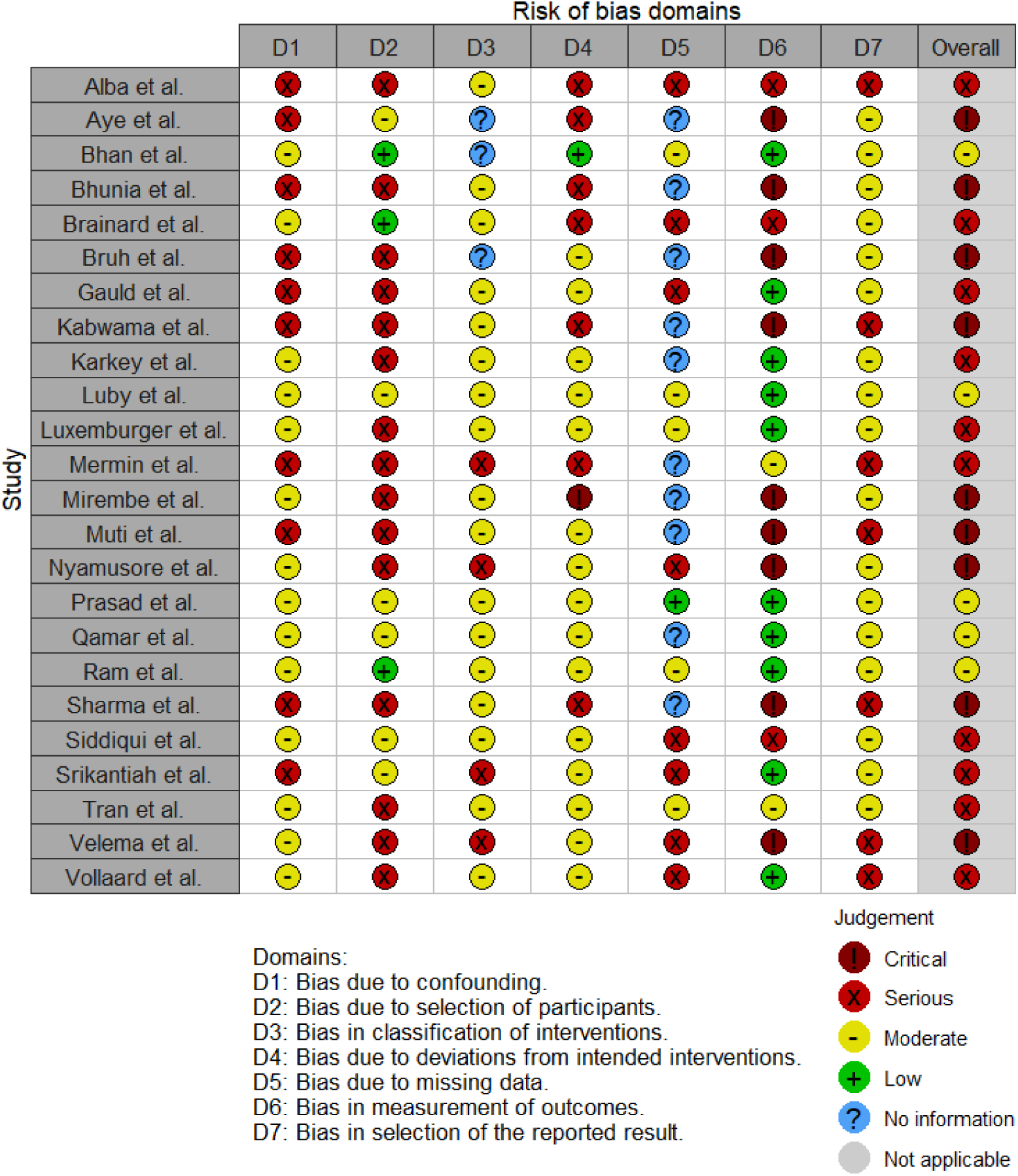
Risk of bias assessment using the Cochrane ROBINS-I tool. The studies included in the systematic review were assessed for risk of bias due to 1) confounding, 2) selection, 3) intervention classification, 4) intervention deviation, 5) missing data, 6) outcome measurement, and 7) selective reporting.

### Water source

JMP definition of improved water source includes piped water, protected dug wells, tube wells, protected springs, rainwater, and packaged water. While the improved water source can be further divided using the service ladders (i.e, safely managed, basic, or limited), we only used one category of improved water source because the number of studies is small and descriptions about the exposure were not detailed enough for further classification. Three studies reported data on the improved water source. The synthesized effect estimate for the odds ratio (OR) of having access to an improved water source was 0.54 with a 95% credible interval (CrI) of 0.31 to 1.08 with the between-study heterogeneity of τ = 0.29.

Drinking water from an unimproved water source (unprotected dug well or spring) or directly from surface water are risk factors for typhoid fever. Five values were fit into the surface water group. Surface water sources increased more than double the odds of having typhoid fever (OR = 2.16, 95% Crl = 1.24 - 3.60, τ = 0.27) (**Figure 3**).

**Figure 3.**
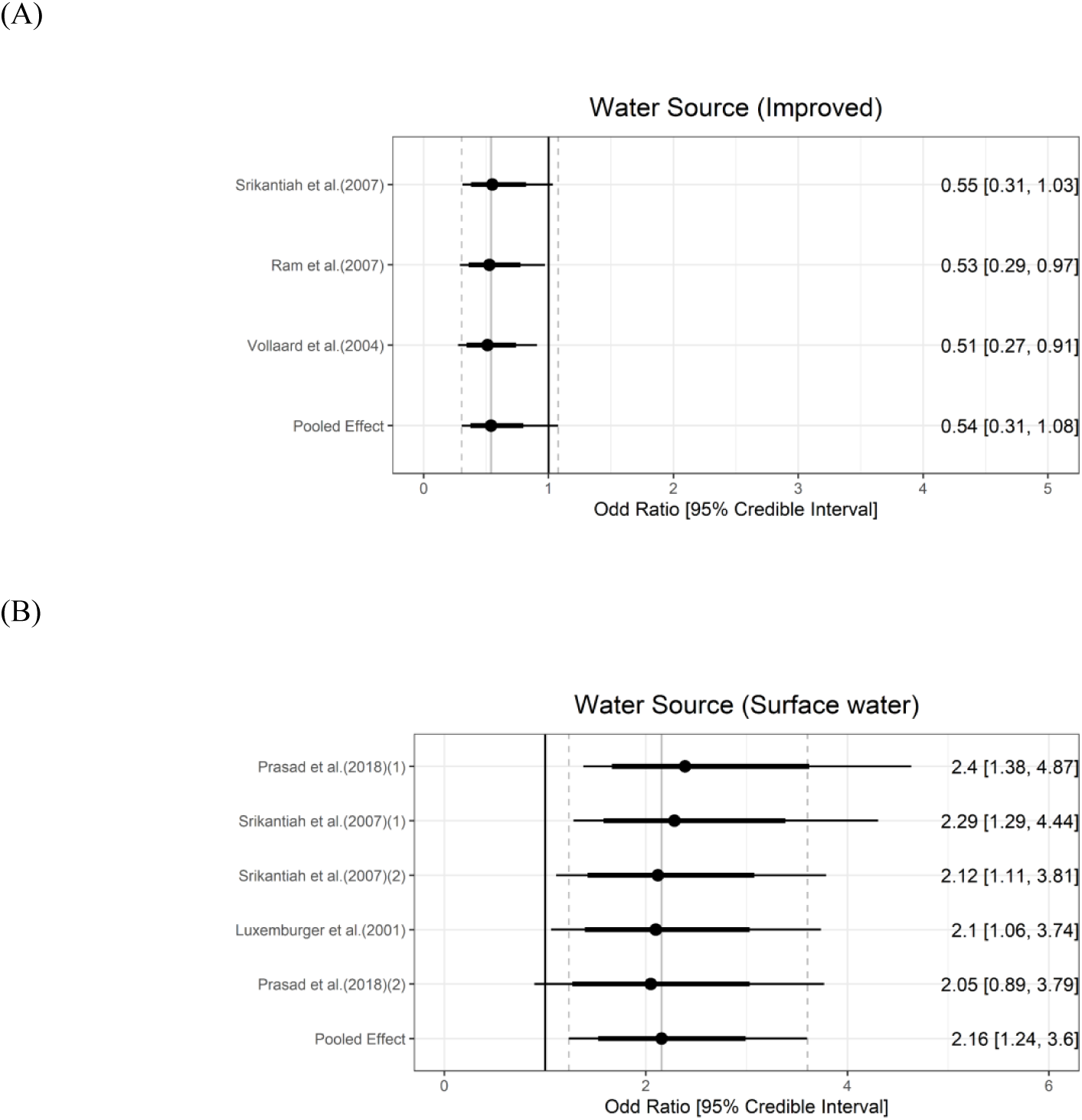
Association between water source and typhoid fever. The forest plot illustrates the association between water source and typhoid fever. Filled circles are posterior median values. Thick and thin black lines show 80% and 95% credible intervals, respectively.

### Water treatment

Household water treatment of any kind was included as a predicted protective factor due to prior evidence on decreasing typhoid fever burden (39). Five studies reported information on water treatment and six exposures were classified as the water treatment group. The meta-analysis showed that any kind of household water treatment was associated with 38% lower odds of typhoid (OR = 0.62, 95% Crl = 0.41 - 0.89, τ = 0.29). Using untreated water (n = 9) was a risk factor and increased the odds of typhoid fever more than double (OR = 2.21, 95% Crl = 1.53 - 3.48, τ = 0.42) (**Figure 4**).

**Figure 4.**
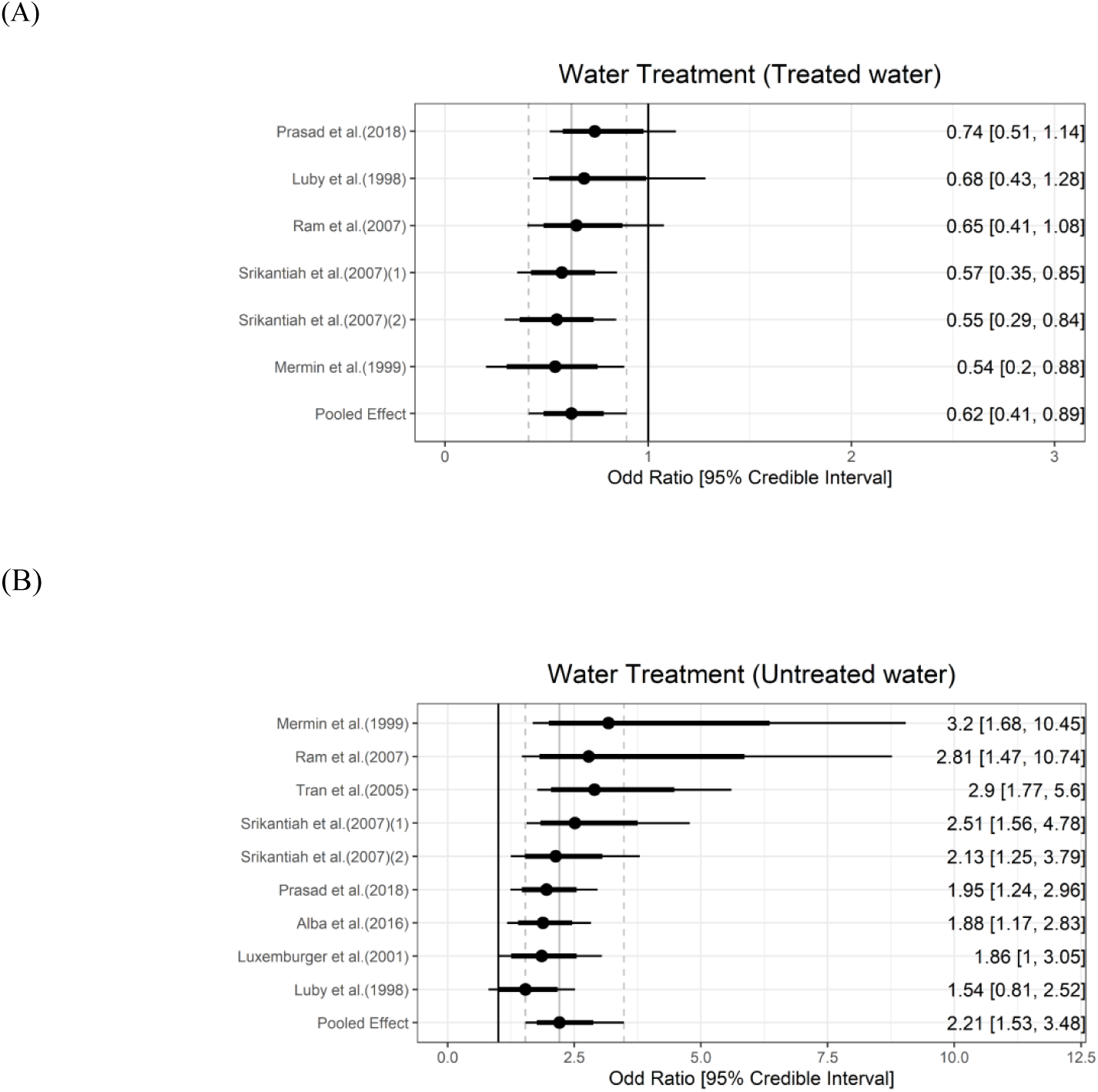
Association between water treatment and typhoid fever. The forest plot illustrates the association between water treatment and typhoid fever. Filled circles are posterior median values. Thick and thin black lines show 80% and 95% credible intervals, respectively.

### Water management

Safely managed water refers to water being stored in a narrow-mouthed, closed lid to prevent contamination (40), and is considered a protective factor against water-borne diseases. In order to expand the concept of safe water management and get a broader pool of data, we considered narrow-mouthed and/or closed lids in our exposure categories. Two studies measured the association between safely managed water and typhoid fever (26,35). Using metal coverage of water storage and keeping water containers covered were associated with around 80% lower odds of having typhoid fever (odds ratio [OR]: 0.22, 95% confidence interval [95% CI]: 0.1-0.6; OR: 0.2, 95% CI: 0.04-1.1) (26,35). Unsafe water management, such as the use of contaminated water storage, is a risk factor, and using dirty containers to store drinking water was associated with double the odds of having typhoid fever (aOR: 1.99, 95% CI: 0.6-6.65) (36). Meta-analysis was not performed in the water management category due to less than three studies.

### Sanitation

JMP defines improved sanitation facilities as those that prevent human contact with excreta. The categories of improved sanitation facilities can be further divided into safely managed, basic, and limited categories. No exposure categories from studies could be classified into these ladder rungs. Prasad et al. (21) measured that people who were using unimproved pit latrine had nearly 50 times greater odds of having typhoid than the controls (aOR: 49.47, 95% CI: 9.42-259.92). On the other hand, open defecation was not significantly associated with odds of typhoid fever (OR = 1.06, 95% Crl = 0.71 - 2.10, τ = 0.39) (**Figure 5**).

**Figure 5.**
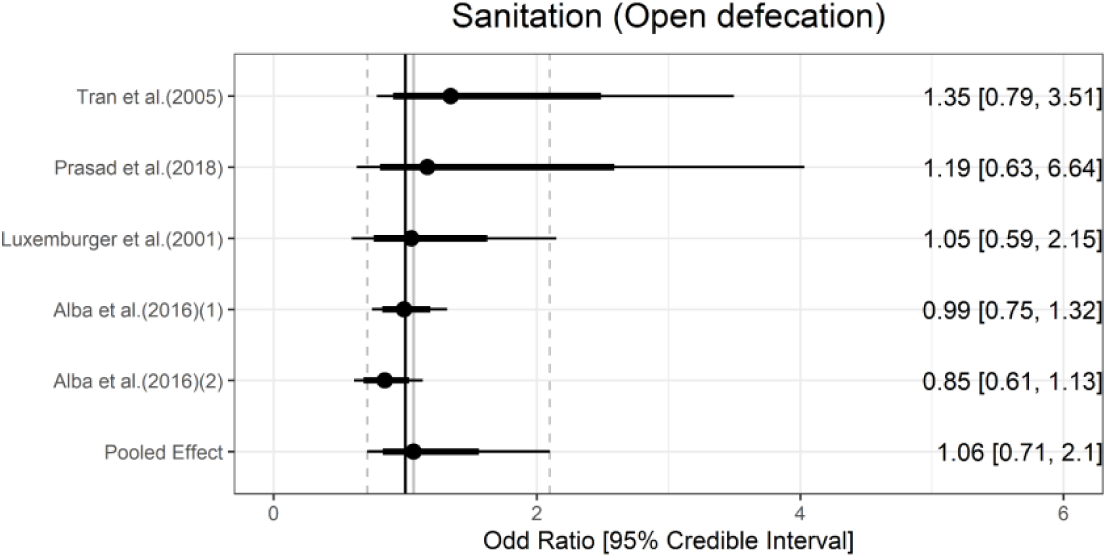
Association between sanitation and typhoid fever. The forest plot illustrates the association between sanitation and typhoid fever. Filled circles are posterior median values. Thick and thin black lines show 80% and 95% credible intervals, respectively.

### Hygiene

According to the JMP definitions, basic hygiene means that a handwashing facility with soap and water is available at home, and washing hands with soap is protective against diarrhea (39). Basic hygiene was significantly associated with 40% lower odds of typhoid fever (OR = 0.60, 95% Crl = 0.38 - 0.97, τ = 0.24). Limited hygiene means that a handwashing facility is available at home without soap and/or water. Limited hygiene was associated with more than double the odds of typhoid fever (OR = 2.26, 95% Crl = 1.38 - 3.64, τ = 0.29) (**Figure 6**).

**Figure 6.**
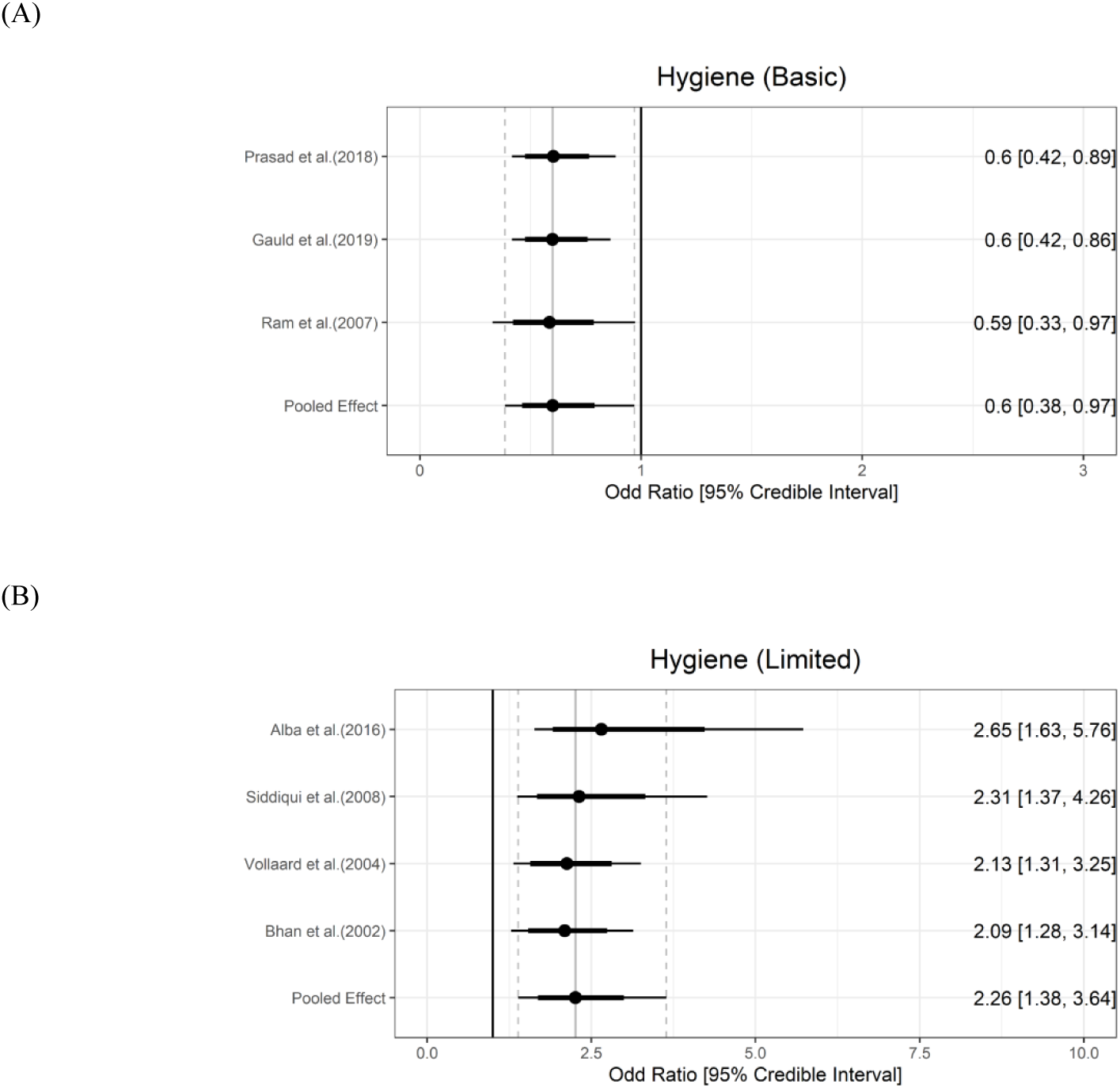
Association between hygiene and typhoid fever. The forest plot illustrates the association between hygiene and typhoid fever. Filled circles are posterior median values. Thick and thin black lines show 80% and 95% credible intervals, respectively.

## Discussion

We have conducted a systematic review and meta-analysis of case-control studies to infer the association between water, sanitation, and hygiene (WASH) and culture-confirmed typhoid fever. Our analyses updated the previous estimates for the association and confirm that improved WASH such as treated water and basic hygiene reduced the odds of typhoid fever. As in the previous review by Brockett *et al*. (11), our analyses indicate that open defecation is not significantly associated with the odds of typhoid fever. We think that even if open defecation increases the transmission of typhoid fever in the community, individuals who reported open defecation do not necessarily have higher odds of typhoid fever if cases and controls are from the same community.

Our analyses updated the previous review by Brockett *et al*. (11) by including new studies identified through a literature search. Also, unlike the previous study, we assessed the risk of bias using the ROBINS-I tool, which was specifically designed to address weaknesses in prior tools in assessing the risk of bias in non-randomized studies (15). Conducting arguably more rigorous risk of bias assessment and identifying potential mistakes in the previous review resulted in a smaller number of studies included in the meta-analysis. We adopted the Bayesian meta-analysis as it was claimed to have advantages in characterizing the uncertainty of the estimates, particularly when the number of studies is small (17) while the difference from the frequentist approach seems minimal in our particular case (**Appendix E**).

While the previous review by Brockett *et al*. (11) included studies in which typhoid fever was confirmed through the Widal test or clinical signs as well as blood culture, we included only studies in which typhoid fever was confirmed through blood culture. Clinical symptoms of typhoid fever are generally not specific enough to differentiate from other diseases and Widal tests are known to have a high false-positive rate (12). We also excluded some estimates that were included in the review for other reasons. For instance, the authors of the previous review included estimates for both sometimes treating water before drinking (i.e., sometimes vs. always) and never treating water before drinking (i.e., never vs. always) from Alba *et al*. (33) as inputs for meta-analysis of the untreated water category, while we only included the later one. Also, the previous review included both crude and adjusted estimates of the same exposure from the same study in the meta-analysis, which we believe violates the assumption of independent findings (i.e., unit-of-analysis error) (41). In this case, we included only adjusted OR estimates in the meta-analysis. Also, when there are multiple exposures that can be classified into the same JMP WASH category (e.g., use of soap and soap near the toilet can be classified into the hygiene category), the previous review included both of the exposure categories whereas we included one of the two exposures that fits the JMP definition better (i.e., soap near the toilet in this case) in the analyses. Also, we utilized more detailed WASH subcategories that were not used in the previous review. Accordingly, some exposures included in the previous meta-analyses were not included in our meta-analysis. For instance, although washing hands before meals regularly or after using the toilet was included in the lack of hygiene category in the previous review, we did not classify these exposures into any of our hygiene categories as washing hands does not imply washing hands with soap, which fits the JMP hygiene category (37).

Our study has several limitations. First, studies that were included in our meta-analyses vary not only in terms of study place and time, but also in how potential biases were controlled in the case-control studies. These variances may not be fully captured by the random-effects model approach. However, the heterogeneities of the OR estimates did not appear to be very high with the highested *I*^2^ statistic being 61.87% for open defecation category. Second, there were discrepancies across studies in how the WASH exposure data was collected. In some studies, the data collection relied on self-reporting (e.g., washing hands with soap) (20,23), whereas in other studies the data was collected through the researcher’s direct observation (e.g., observation of soap availability) (24,36,37). Third, various WASH indicators may be related to the habits of an individual and thus correlated with one another. This implies that some of the included studies that do not control for other WASH factors can not differentiate the impacts of different WASH components. Fourth, while we used our best judgment to categorize the WASH exposures in case-control studies according to JMP categories, actual WASH exposures from one JMP WASH category still vary. Lastly, we only included findings from case-control studies as we are updating the previous review of case-control studies. Findings from randomized controlled trials (42,43) and cohort studies (44) are consistent with our analyses. For example, in a clinical trial conducted in Kolkata, India, living in a better WASH environment led to 57% (95% CI: 15 - 78) reduction in typhoid risk (42).

There are opportunities for future research. There were only a few or no included studies for some JMP WASH categories (e.g., unimproved water source, safely managed sanitation, basic sanitation, limited sanitation, and no hygiene facility), for which additional studies will enrich our understanding between WASH and typhoid fever. Our findings, when combined with population-level JMP WASH trends, may be used to understand and forecast the population-level risk of typhoid fever, which can provide essential insights for decision-makers. Since the population levels of WASH have been monitored since 1990 in 191 countries, one can also analyse the longitudinal data to explore the country-level association and longitudinal trends between the levels of WASH and typhoid fever burden.

Our study findings will be useful to infer actionable insights on the most effective ways to control typhoid fever in LMICs. For instance, our findings reinforce the previous findings that, in addition to infrastructure improvements, behavioural changes such as washing hands with soap can have a significant impact on the risk of contracting typhoid fever (45). While major infrastructural improvements are crucial to reduce the burden of typhoid fever, they require resources that is difficult to commit to in LMICs. On the other hand, behavior interventions may be feasible, affordable, and effective options to reduce the disease risk in LMICs. In conclusion, improving WASH through improved water source, water treatment, and basic hygiene was associated with a reduced risk of typhoid fever and should remain a cornerstone for the prevention and control of typhoid fever.

## Role of the funding source

This work was supported, in whole or in part, by Gavi, the Vaccine Alliance, Bowdoin College, and the Bill & Melinda Gates Foundation, via the Vaccine Impact Modelling Consortium (Grant Number OPP1157270 / INV-009125). The funders were not involved in the study design, data analysis, data interpretation, and writing of the manuscript. The authors alone are responsible for the views expressed in this article and they do not necessarily represent the decisions, policy, or views of their affiliated organisations.

## Contributions

JHK conceptualised and designed the study. GG and CK extracted data from the included studies. CK and JHK examined the risk of bias and conducted the meta-analyses using statistical software. CK, GG, JHK wrote the first draft, and all authors contributed to reviewing and editing the manuscript and have approved the final version.

## Data Availability

All data produced are available online at https://github.com/ckim0509/WASH_Typhoid

## Acknowledgements

We thank Justin Im (International Vaccine Institute) and John D. Clemens (International Vaccine Institute) for their review and feedback on this article.

## Declaration of interests

We declare no conflict of interest.

## Notes

### Competing Interest Statement

The authors have declared no competing interest.

