## Supplementary appendix for "Associations of Water, Sanitation, and Hygiene with Typhoid Fever in Case-Control Studies: A Systematic Review and Meta-Analysis"

**Appendix A. Data extracted from the included studies** This file contains information we extracted from the included papers. This was used as an input for the statistical analysis. (Author = author name; Year = publication year; Study = year and author, Country = country,  Exposures =  details of exposure, Measures = level of exposure; JMP WASH Category = classified category using JMP WASH category; Brockett Category = category used in the previous review; Crude OR = odds ratio from univariate analysis; Crude OR CI = confidence interval of odds ratio from univariate analysis; Adjusted OR = odds ratio from multivariate analysis; Adjusted OR CI = confidence interval of odds ratio from multivariate analysis; Diagnostic Methods = diagnostic methods used to define typhoid fever; Blood Culture = Blood culture-based diagnosis of typhoid)

Download (download cvs file)

**Appendix B. Funnel plots** To assess publication bias, we evaluated funnel plot asymmetry. Significant funnel plot asymmetry was not detected in our analyses.

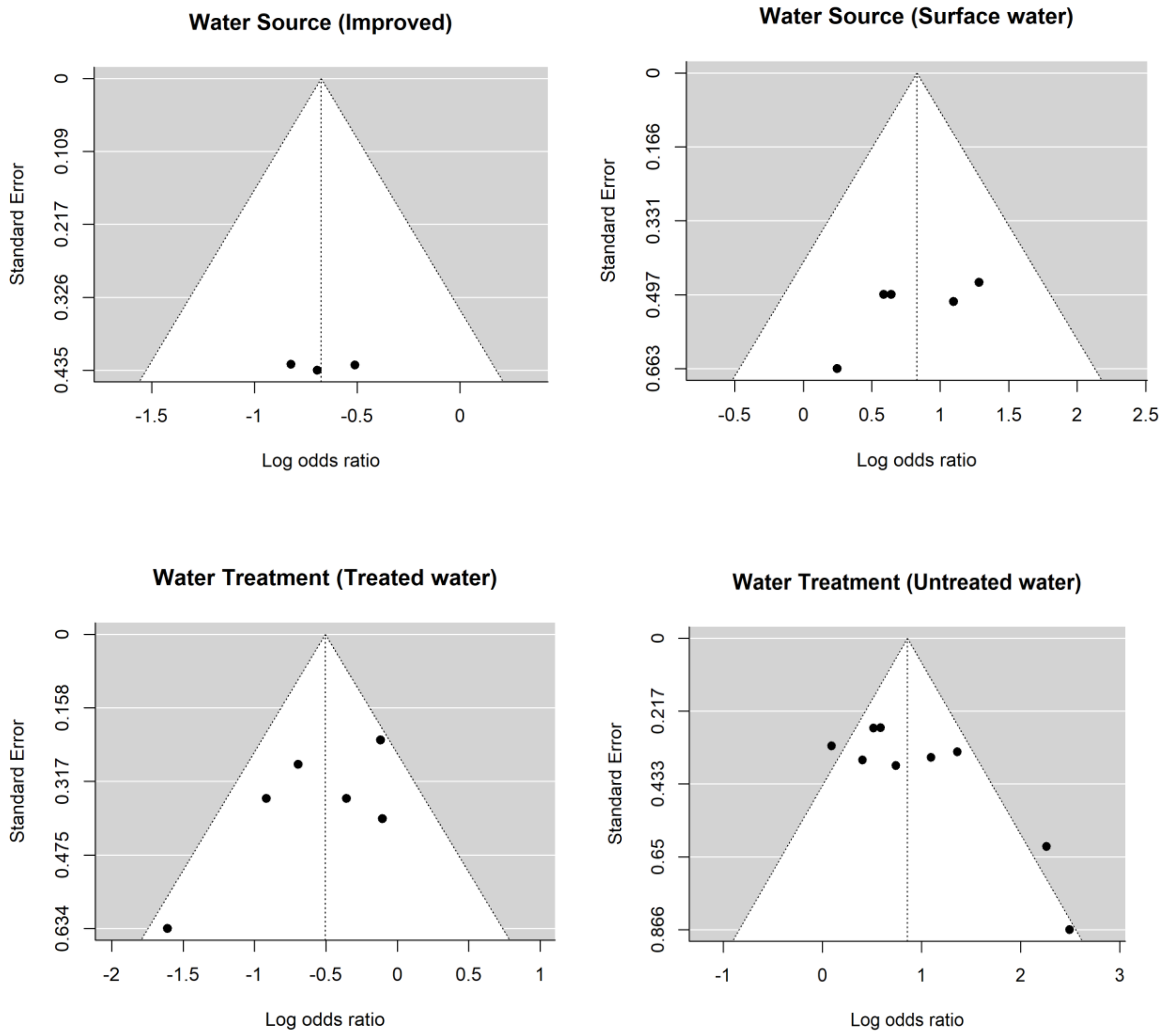

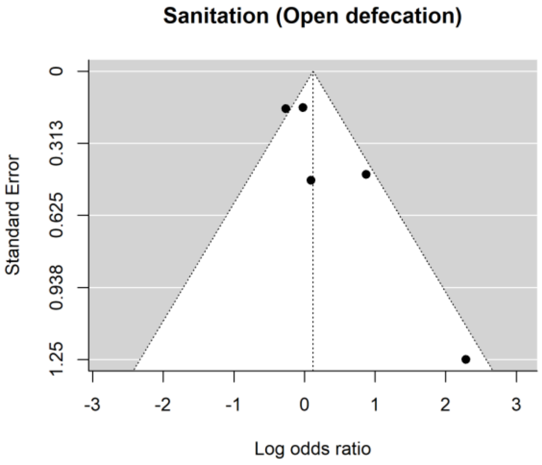

Sanitation (Open defecation)
Standard Error
0
0.313
0.625
0.938
1.25
-3
-2
-1
0
1
2
3
Log odds ratio


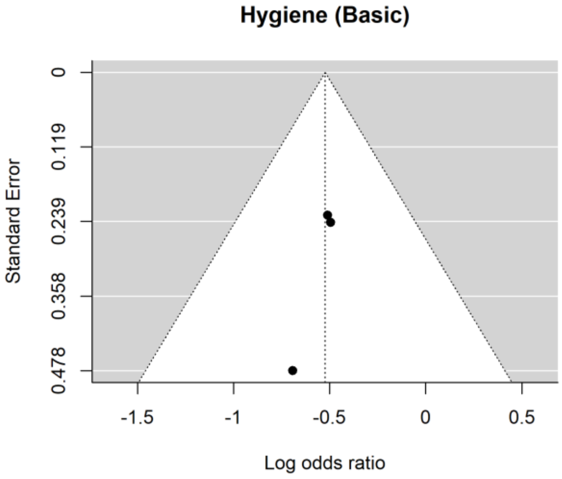

Hygiene (Basic)
Standard Error
0
0.119
0.239
0.358
0.478
-1.5
-1
-0.5
0
0.5
Log odds ratio


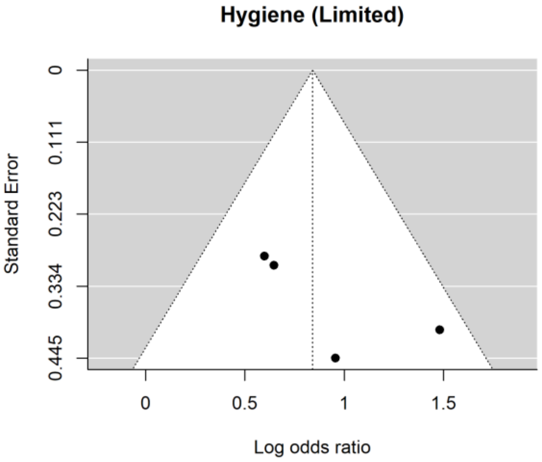

Hygiene (Limited)
Standard Error
0
0.111
0.223
0.334
0.445
0
0.5
1
1.5
Log odds ratio

**Appendix C. Risk of bias assessment results** This is risk of bias assessment results broken down for each risk of bias criterion.

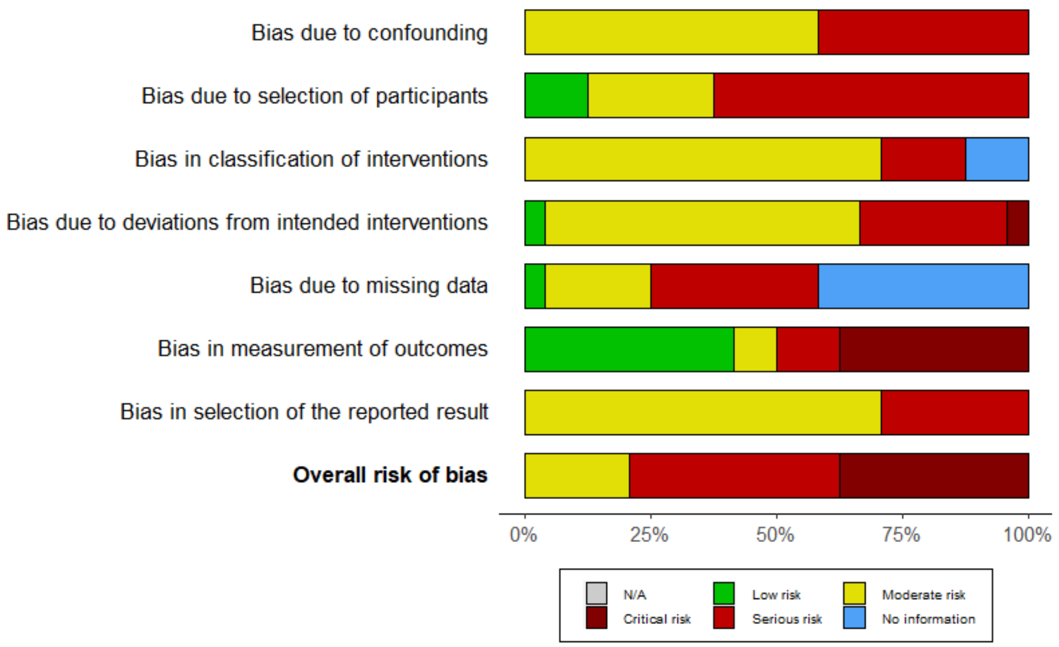

**Appendix D. Model description** This model describes the Bayesian random effects model used in our study

As below, observation, $y_i$ (i.e., the log(ORs) from the study $i$) is assumed to be normally distributed for a given mean $\theta_i$ and standard error $\sigma_i$. Here, $\theta_i$ is again assumed to be a normal random variable with a mean $\mu$ and variance $\tau^2$.

$$y_i \sim \text{Normal}(\theta_i, \sigma_i^2)$$

$$\theta_i \sim \text{Normal}(\mu, \tau^2)$$

$$\mu \sim \text{Normal}(0,1)$$

$$\tau \sim \text{Half-Cauchy}(0,0.5)$$

Our main interest is to estimate the true pooled effect size $\mu$ and the between-study heterogeneity , $\tau$ . We defined prior distribution of μ as a normal distribution and τ as a half-Cauchy distribution with the location parameter of 0 and the scaling parameter of 0.5. The overall convergence and validity of the model were confirmed using the potential scale reduction factor and posterior predictive checks. Based on the assessments, we used 10,000 iterations of the MCMC algorithm.

**Appendix E. Summaries of the meta-analyses using Frequentist and Bayesian Meta-Analysis** This table shows the pooled estimates using both frequentist and Bayesian approaches. Heterogeneity in frequentist meta-analysis was assessed by using the $I^2$ statistic.

| WASH indicators | *N** | Pooled OR [95% CrI]† | Pooled OR [95% CI]†† | $I^2$ |
|---|---|---|---|---|
| Improved water source | 4 | 0.54 [0.31, 1.08] | 0.51 [0.31, 0.83] | 0.00 % |
| Treated Water | 6 | 0.62 [0.41, 0.89] | 0.60 [0.43, 0.85] | 37.41% |
| Basic hygiene | 3 | 0.60 [0.38, 0.97] | 0.59 [0.44, 0.81] | 0.00% |
| Surface water | 5 | 2.16 [1.24, 3.60] | 2.29 [1.46, 3.61] | 0.00 % |
| Untreated water | 9 | 2.21 [1.53, 3.48] | 2.36 [1.60, 3.49] | 61.81% |
| Open defecation | 5 | 1.06 [0.71, 2.10] | 1.13 [0.73, 1.75] | 61.87% |
| Limited hygiene | 4 | 2.26 [1.38, 3.64] | 2.32 [1.60, 3.36] | 15.95% |

* Number of exposures
† Pooled estimates using Bayesian meta-analysis approach
†† Pooled estimates using frequentist meta-analysis approach
